# Exposure to angiotensin-converting enzyme inhibitors that cross the blood-brain barrier and the risk of dementia among patients with human immunodeficiency virus

**DOI:** 10.1101/2024.01.16.24301275

**Authors:** Tammy H. Cummings, Joseph Magagnoli, Aliaksandra Sikirzhytskaya, Ilya Tyagin, Ilya Safro, Michael D. Wyatt, Michael Shtutman, S. Scott Sutton

## Abstract

More than one million people in the United States and over 38 million people worldwide are living with human immunodeficiency virus (HIV) infection. Antiretroviral therapy (ART) greatly improves the health of people living with HIV (PLWH); however, the increased life longevity of PLWH has revealed consequences of HIV-associated comorbidities. HIV can enter the brain and cause inflammation even in individuals with well-controlled HIV infection. The quality of life for PLWH can be compromised by cognitive deficits and memory loss, termed HIV-associated neurological disorders (HAND). HIV-associated dementia is a related but distinct diagnosis. Common causes of dementia in PLWH are similar to the general population and can affect cognition. There is an urgent need to identify treatments for the aging PWLH population. We previously developed AI-based biomedical literature mining systems to uncover a potential novel connection between HAND the renin-angiotensin system (RAAS), which is a pharmacological target for hypertension. RAAS-targeting anti-hypertensives are gaining attention for their protective benefits in several neurocognitive disorders. To our knowledge, the effect of RAAS-targeting drugs on the cognition of PLWH development of dementia has not previously been analyzed. We hypothesized that exposure to angiotensin-converting enzyme inhibitors (ACEi) that cross the blood brain barrier (BBB) reduces the risk/occurrence of dementia in PLWH. We report a retrospective cohort study of electronic health records (EHRs) to examine the proposed hypothesis using data from the United States Department of Veterans Affairs, in which a primary outcome of dementia was measured in controlled cohorts of patients exposed to BBB-penetrant ACEi versus those unexposed to BBB-penetrant ACEi. The results reveal a statistically significant reduction in dementia diagnosis for PLWH exposed to BBB-penetrant ACEi. These results suggest there is a potential protective effect of BBB ACE inhibitor exposure against dementia in PLWH that warrants further investigation.

## 1.0 Introduction

Human Immunodeficiency Virus (HIV) is a viral infection that weakens the immune system leading to the development of HIV associated comorbidities (e.g., opportunistic infections). More than 38 million people worldwide have a diagnosis of HIV-1 infection, and more than 1.2 million are in the United States.^1^ Fortunately, antiretroviral therapy (ART) is highly effective at managing HIV by suppressing the replication of HIV reducing the viral load to undetectable levels, improving overall health and quality of life for people living with HIV (PLWH). The increased longevity of PLWH has also unfortunately revealed longer-term consequences of HIV-associated comorbidities. HIV can enter the brain early in the course of an infection, which establishes a reservoir and causes persistent inflammation and cognitive deficits. ART does not cross the blood brain barrier (BBB) and subsequently the central nervous system (CNS) remains a viral sanctuary and causes neurological damage for PLWH.^2^ People living with HIV are at an increased risk of experiencing cognitive issues, including mild cognitive impairment and HIV-associated neurocognitive disorders (HAND). Additionally, there is a link between HIV and other forms of cognitive decline (e.g., dementia). HAND and dementia are related but distinct conditions. HAND specifically refers to cognitive impairment that is directly linked to the effects of HIV on the central nervous system. Dementia in individuals with HIV refers to cognitive impairment that results from various causes that can be unrelated to the direct effects of HIV on the brain. Causes of dementia in PLWH are similar to the general population and include age-related factors, cardiovascular disease, and comorbidities that can affect cognition. Consequent to the effectiveness of ART, individuals 50 years or greater of age account for more than 50% of HIV-1 seropositive individuals in high-resource countries^3^; a prevalence that is expected to reach approximately 73% by 2030.^4^ Therefore, there is a critical urgency to discover therapeutics to prevent or delay the neurocognitive decline of the aging PLWH population.

Biomedical data and literature are exponentially growing, making it difficult to analyze and discover hidden connections in these vast datasets. Therefore, we developed an artificial intelligence (AI)-based literature mining system that discovers novel interactions and applied them to discovering drugs that can be repurposed for the management of cognitive decline.^5–8^ The AI systems prioritize mining results to uncover small molecules that can be repurposed for new therapies, including drugs already approved by the FDA for other indications. Several molecules identified by these AI systems have been experimentally validated as neuroprotective agents in a model of HIV-associated neuronal damage exacerbated by drugs of abuse.^7,9^ We developed and employed an AI system called Automatic Graph-mining And Transformer based Hypothesis generation Approach (AGATHA) to uncover a potential novel connection between HAND and the renin-angiotensin system (RAAS), which is well known for its role in vasoconstriction and is also a pharmacological target in the treatment of hypertension.^10^

To our knowledge, the effect of ACE inhibitors on the cognition of PLWH development of dementia has not previously been analyzed, yet we posit that the experiments have already been performed. Electronic Health Records (EHRs) contain patient health information, which when analyzed in aggregate provides opportunities to examine entire populations by classifying patient phenotypes. Such patient phenotypes may be the most accurate means of representing the interplay of all the health determinants of precision medicine: genes, environment and lifestyle.^11^ Our team has a demonstrated success record in such retrospective analyses. We hypothesized that exposure to ACE inhibitors that cross the BBB reduces the risk/occurrence of dementia in PLWH and report a retrospective cohort study of EHRs to examine the proposed hypothesis.

## 2.0 Methods

### 2.1 Text mining

Previously we developed and employed Automatic Graph-mining And Transformer based Hypothesis generation Approach (AGATHA), which is a deep-learning hypothesis generation system that learns a data driven ranking criteria to recommend new biomedical connections. AGATHA employs a range of natural language processing (NLP) techniques to generate hypotheses linking specific terms provided to the system and ranks the hypotheses based on their likelihood and potential significance.^5–8^ We applied AGATHA to determine pathways and drugs with potential to be repurposed for the treatment of HIV-associated neurocognitive disorder (HAND). The connection of all human genes with HAND was tested and ranked as previously done in Aksenova et al.^7^ The high-ranked genes and pathways targeted by FDA-approved drugs were selected for the future analysis. Among the most promising pathways was the renin-angiotensin pathway and specifically ACEi inhibitors. The AGATHA system is publicly available at https://github.com/IlyaTyagin/AGATHA-C-GP, 2021.^8^

### 2.1 Data & Data Source

This retrospective cohort drug-disease association study was conducted using data from the United States Department of Veterans Affairs. The VA Informatics and Computing Infrastructure (VINCI) was utilized to obtain individual-level information on demographics, administrative claims, and pharmacy dispensation. The completeness, utility, accuracy, validity, and access methods are described on the VA website, https://www.hsrd.research.va.gov/for_researchers/vinci/. The study was conducted in compliance with the Department of Veterans Affairs requirements and received Institutional Review Board and Research and Development Approval. The study utilized inpatient and outpatient claims coded with International Classification of Diseases (ICD) revision 9-CM, revision 10-CM, Current Procedure Terminology (CPT), pharmacy, laboratory, and vital sign data.

### 2.2 Cohort Selection

Patients with an ICD 9/10 diagnosis code of human immunodeficiency virus (HIV) and hypertension were included in the study. Patients with HIV were extracted from the outpatient and inpatient claims files and defined as ICD-9CM code of 042.x, V08, V65.44 or ICD-10CM code of B20.x-B24.x or Z21.x. Among the cohort of PLWH, patients with hypertension were identified by extraction from the outpatient and inpatient claims files using, ICD-9CM 401.x, 402.x, 403.x, 404.x, 405.x or ICD-10CM I10, I11.x, I12.x, I13.xx, I15.x, I16.x with their first hypertension diagnosis (study index) between October 2000 and March 31, 2022. Inclusion criteria for the study included: 1) HIV diagnosis before hypertension diagnosis; 2) no blood brain barrier (BBB) angiotensin converting enzyme (ACE) inhibitor prescription prior to initial hypertensive diagnosis; and 3) at least 1 year of follow-up after study index. Exclusion criteria for the study included: 1) a diagnosis of dementia prior to the study index or 2) prescription of BBB ACE inhibitor prior to study index. Patients were followed from study index until the first date of a) dementia diagnosis, b) death, or c) March 31, 2023. *In a sub-analysis*, we have included similar inclusion/exclusion criteria to the cohort described. Inclusion criteria included: 1) HIV diagnosis before hypertension diagnosis; 2) no ACE inhibitor prescription prior to initial hypertensive diagnosis; and 3) at least 1 year of follow-up after study index. Exclusion criteria included: 1) a diagnosis of dementia prior to the study index; 2) prescription of any ACE inhibitor prior to study index; or 3) prescriptions for both BBB ACE inhibitor and non-BBB ACE inhibitor during the study period.

### 2.3 Study Outcome

The study outcome was incident dementia. A diagnosis of dementia was identified from inpatient and outpatient claims data via ICD-9CM (290.x, 294.1, 294.2, 331.0, 331.1, 331.2, 331.82) and ICD-10CM (F00.x, F01.x, F02.x, F03.x, G30.x) codes.

### 2.4 Medication Exposure

Patients were grouped into two mutually exclusive cohorts based on any exposure to BBB ACE inhibitor(s) between study index and the end of study. BBB ACE inhibitors consisted of captopril, fosinopril, lisinopril, perindopril, ramipril, and trandolapril and non-BBB ACE inhibitors included: benazepril, enalapril, moexipril, and quinapril.^12,13^ Therefore, exposure groups were defined as: (a) BBB ACEi exposed; and (b) BBB ACEi unexposed. In the sub-analysis, we also compare those exposed to a BBB ACEi versus a non-BBB ACEi.

### 2.5 Baseline Data

Demographic and clinical characteristics included age, sex, race/ethnicity, body mass index (BMI), and year of study index. Comorbid conditions include all conditions included in the Charlson Comorbidity Index (CCI), current smoking (VA HealthFactor dataset), hypercholesterolemia, hypertriglyceridemia, hyperlipidemia, ischemic heart disease, other heart disease, type 2 diabetes, atrial fibrillation, hypothyroidism, hyperthyroidism, depression, traumatic brain injury, alcohol dependence, Parkinson’s disease, generalized anxiety disorder, and chronic kidney disease.

### 2.6 Statistical Analysis

Baseline cohort characteristics were compared with statistical tests, including t-tests and chi-square tests. Differences among the cohorts were also quantified using the standardized difference, which is relatively invariant to sample size compared to the p-value. Cox proportional hazards models, unadjusted and adjusted for all covariates, were fit and hazard ratios (HR) with corresponding 95% confidence intervals (CIs) are presented. A propensity score model included all demographic, clinical, and comorbidity as predictors of BBB ACE treatment. Differences were again quantified by using statistical tests such as t-tests, chi-square test and standardized difference. A Cox proportional hazard model was used to analyze the risk of dementia for matched sample. To evaluate the robustness of our findings we calculate the E-values for the outcome model. The E-value is the minimum association required for an unmeasured confounder to have with both the treatment and outcome to nullify the treatment effect.^14,15^ In the sub-analysis, we analyze differences between the ACE inhibitor exposed cohorts similar to original analyses above, and provide a univariate Cox proportional hazard model after a propensity score match with all covariates, while exact matching on those variables that were statistically significant between the cohorts (race, BMI, and hypertriglyceridemia). Data management and analysis was performed using SAS (SAS Institute Inc., SAS Enterprise Guide 8.3, Cary, NC: SAS Institute Inc.).

## 3.0 Results

We previously applied AGATHA to seek potential drugs to be repurposed for the treatment of HIV-associated neurocognitive disorder (HAND), during which the connection of all human genes with HAND were ranked (REF). The highest-ranked genes and pathways for which FDA-approved drugs are known were prioritized for detailed analysis. From this prioritized list, several small molecules have been experimentally validated with in vitro models.^7,9,16^ Further analysis of high-ranking hits revealed the renin-angiotensin pathway and specifically ACE inhibitors. The target of the most common antihypertension drugs (ACE inhibitors (ACEi), and ARBs: angiotensin-2 receptor 1 blockers)^17,18^ coincidentally include some drugs that are capable of crossing the blood brain barrier (BBB) and attention is growing for their potential positive effects on cognition. For example, recent retrospective meta-analysis studies show that the BBB-crossing ACEi and ARBs improve the cognition of the elderly and delay Alzheimer’s Disease progression.^12,13,19^ Common causes of dementia in PLWH are similar to the general population and include age-related factors, cardiovascular disease, and other comorbidities that can affect cognition. For example, hypertension is a comorbidity that is a well-established risk factor for cognitive decline and dementia in the general population.^20^ Hypertension can cause deficits in memory, function, attention, and psychomotor speed compared to normotensive individuals.^21–24^ Interestingly, patients with hypertension receiving antihypertensives had a decreased risk of dementia and medications targeting the RAAS have been reported to confer the greatest benefit, but the results have been discordant.^25–32^. We therefore set out to examine this potential connection with a retrospective analysis of health records.

A total of 18,250 patients met the inclusion and exclusion criteria and were evaluated in our study. The cohorts consisted of 9,419 patients exposed to an BBB ACE inhibitor (ACEi) and 8,831 patients unexposed to a BBB ACEi. The mean age was lower in the BBB ACEi exposure group (51.95 years, SD=9.4) compared to the BBB ACEi unexposed group (53.09 years, SD=10.41) (p<0.001). A higher proportion of females were found in the group BBB ACEi unexposed group (3.68%) compared to the group with exposure to BBB ACEi (2.45%) (p<0.001, standardized difference=0.071). However, the majority of individuals in both cohorts were male, with a higher percentage in the BBB ACEi exposure group (97.55%) than in the BBB ACEi unexposed group (96.32%). Regarding race, in the BBB ACEi exposure group, 51.06% were Black and 43.37% were White. In comparison, the BBB ACEi unexposed group had 48.87% Black individuals and 43.03% White individuals. For BMI (Body Mass Index), there were notable differences in distribution. In the BBB ACEi exposure group, 2% had a BMI of less than 18.5, 33.8% had a BMI between 18.5 and 24.9, 38.75% had a BMI between 25.0 and 29.9, and 25.29% had a BMI of 30 or higher. The ACEi exposed group had 3.18%, 36.67%, 36.43%, and 22.98% in the respective BMI categories. There was a statistically significant difference among the Charlson comorbidity index (CCI); however, the cohorts had a CCI of 4.71 (BBB ACEi) and 5.23 (BBB ACEi unexposed) indicating a high percentage of comorbidities among both groups. Additional baseline demographics and clinical characteristics and standard differences among the cohorts are included in Table 1.

**Table 1:** Baseline Demographic and Clinical Characteristics.

|  |  | BBB ACE Inhibitor<br>Exposed(N=9419) | BBB ACE<br>Inhibitor<br>Unexposed<br>(N=8831) | p-value | Standardized<br>Difference |
| --- | --- | --- | --- | --- | --- |
| Sex, No (%) | Female | 231(2.45) | 325(3.68) | <0.001 | 0.071 |
|  | Male | 9188(97.55) | 8506(96.32) |  | 0.071 |
| Race, No (%) | Black | 4809(51.06) | 4316(48.87) | <0.001 | 0.044 |
|  | Other/Unknown | 525(5.57) | 715(8.1) |  | 0.100 |
|  | White | 4085(43.37) | 3800(43.03) |  | 0.007 |
| Age, mean(SD) |  | 51.95(9.4) | 53.09(10.41) | <0.001 | 0.115 |
| Charlson comorbidity, mean(SD) |  | 4.71(3.13) | 5.23(3.05) | <0.001 | 0.170 |
| BMI, No (%) | <18.5 | 188(2) | 281(3.18) | <0.001 | 0.074 |
|  | 18.5-24.9 | 3184(33.8) | 3238(36.67) |  | 0.060 |
|  | 25.0-29.9 | 3650(38.75) | 3217(36.43) |  | 0.048 |
|  | 30+ | 2382(25.29) | 2029(22.98) |  | 0.054 |
|  | Missing | 15(0.16) | 66(0.75) |  | 0.088 |
| Pure hypercholesterolemia, No (%) |  | 258(2.74) | 218(2.47) | 0.252 | 0.017 |
| Hypertriglyceridemia, No (%) |  | 382(4.06) | 376(4.26) | 0.494 | 0.010 |
| Hyperlipidemia, No (%) |  | 1324(14.06) | 1573(17.81) | <0.001 | 0.103 |
| Ischemic heart disease, No (%) |  | 391(4.15) | 400(4.53) | 0.210 | 0.019 |
| Other heart disease, No (%) |  | 388(4.12) | 412(4.67) | 0.072 | 0.027 |
| T2DM, No (%) |  | 662(7.03) | 294(3.33) | <0.001 | 0.168 |
| Cerebral infarction, No (%) |  | 14(0.15) | 19(0.22) | 0.291 | 0.016 |
| Atrial fibrillation, No (%) |  | 36(0.38) | 39(0.44) | 0.531 | 0.009 |
| Hypothyroidism, No (%) |  | 92(0.98) | 137(1.55) | 0.001 | 0.051 |
| Hyperthyroidism, No (%) |  | 21(0.22) | 29(0.33) | 0.173 | 0.021 |
| Depression, No (%) |  | 1538(16.33) | 1412(15.99) | 0.534 | 0.009 |
| TBI, No (%) |  | 22(0.23) | 23(0.26) | 0.715 | 0.006 |
| Alcohol dependence, No (%) |  | 769(8.16) | 674(7.63) | 0.183 | 0.020 |
| Parkinson's disease, No (%) |  | ss | 11(0.12) | 0.024 | 0.033 |
| Generalized anxiety disorder, No (%) |  | 115(1.22) | 119(1.35) | 0.448 | 0.012 |
| Chronic kidney disease, No (%) |  | 165(1.75) | 163(1.85) | 0.633 | 0.008 |
| Smoker, No (%) |  | 4649(49.36) | 3898(44.14) | <0.001 | 0.105 |
| Index Year, mean(SD) |  | 2004.3(5.47) | 2006.05(6.41) | <0.001 | 0.294 |

ss= sample size 10 patients or less.

A univariate Cox proportional hazard model demonstrated that patients exposed to BBB ACEi had a 21.4% lower hazard of dementia compared to control BBB ACEi unexposed cohort (HR 0.786, 95% CI 0.692-0.893; Table 2). Furthermore, a Kaplan Meir plot of incident dementia demonstrates that patients exposed to BBB ACEi had a higher probability of disease-free time compared to the control BBB ACEi unexposed cohort (Figure 1, LogRank test = 0.0002). There are many variables that may influence the development of dementia; therefore, a multivariate Cox model was utilized to account for the baseline demographic and clinical characteristics. The multivariate Cox model result was consistent with the univariate results. For BBB ACE inhibitor exposure, the HR was 0.829 (95% CI: 0.727-0.944), indicating that individuals exposed to BBB ACE inhibitors had a 17.1% lower hazard of dementia compared to those unexposed to BBB ACE inhibitor (Table 3). There are variables within the model that were statistically significant and include race, age, and select comorbidities. The E-score for the point estimates was 1.71 for the unmatched samples. The E-score indicates that for the treatment effects to be nullified (equal to 1) an unmeasured confounder would have to be associated with both the treatment and outcome by a hazard ratio of 1.71 for the unmatched sample.

**Figure 1:**
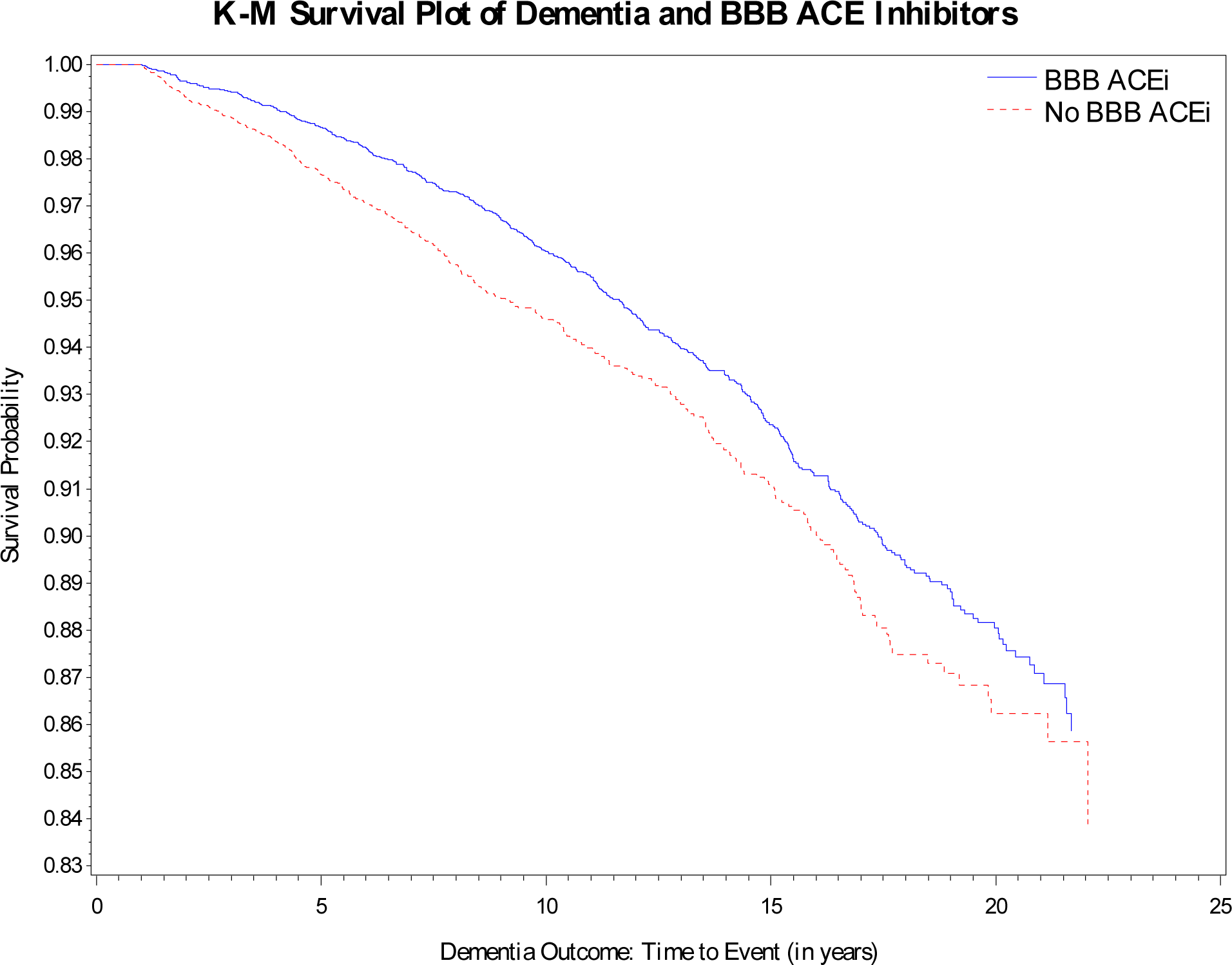
KM curve of original cohort.

**Table 2:** Univariate Results.

|  |  | Cox Model |
| --- | --- | --- |
|  |  | HR (95% CI) |
| BBB ACE Inhibitor Exposure | Yes vs. No | 0.786 (0.692-0.893) |

**Table 3:** Multivariate Statistical Model for Incident Dementia.

|  |  | Cox Model<br>HR (95% CI) |
| --- | --- | --- |
| BBB ACE Inhibitor Exposure | Yes vs. No | 0.829 (0.727-0.944) |
| Sex | Male vs. Female | 0.855 (0.579-1.262) |
| Race | Black vs. White | 1.416 (1.234-1.624) |
|  | Other/Unknown vs. White | 1.043 (0.76-1.432) |
| Age |  | 1.072 (1.064-1.08) |
| Charlson comorbidity |  | 1.013 (0.989-1.039) |
| BMI | <18.5 vs. 18.5-24.9 | 1.297 (0.909-1.852) |
|  | 25.0-29.9 vs. 18.5-24.9 | 0.843 (0.731-0.972) |
|  | 30+ vs. 18.5-24.9 | 0.788 (0.657-0.946) |
|  | Missing vs. 18.5-24.9 | 0.468 (0.116-1.889) |
| Pure hypercholesterolemia |  | 0.646 (0.433-0.964) |
| Hypertriglyceridemia |  | 0.782 (0.549-1.114) |
| Hyperlipidemia |  | 0.976 (0.815-1.168) |
| Ischemic heart disease |  | 1.313 (1.004-1.716) |
| Other heart disease |  | 1.143 (0.859-1.52) |
| T2DM |  | 1.458 (1.159-1.834) |
| Cerebral infarction |  | 2.806 (1.154-6.822) |
| Atrial fibrillation |  | 1.383 (0.68-2.813) |
| Hypothyroidism |  | 1.077 (0.668-1.736) |
| Hyperthyroidism |  | 0.81 (0.256-2.565) |
| Depression |  | 1.316 (1.12-1.547) |
| TBI |  | 2.481 (1.026-5.999) |
| Alcohol dependence |  | 1.221 (0.987-1.51) |
| Parkinson's disease |  | 5.837 (2.362-14.426) |
| Generalized anxiety disorder |  | 0.887 (0.51-1.543) |
| Chronic kidney disease |  | 1.062 (0.685-1.645) |
| Smoker |  | 1.009 (0.882-1.154) |
| Index Year |  | 0.99 (0.975-1.006) |
| E-values: Point Estimate |  | 1.71 |
| Confidence Interval |  | 1.31 |

There are intrinsic limitations to health insurance claims database analyses, including proper documentation, coding, and non-randomized treatment groups. To address these potential limitations, we utilized propensity score matching to assemble cohorts of patients with similar baseline characteristics with the attempt to reduce possible bias in estimating treatment effects. Table 4 lists the baseline characteristics and clinical characteristics of the propensity matched analysis. A total of 15,594 patients were evaluated in the propensity score matching (7,797 in each cohort). Although the standardized differences among the baseline and clinical characteristics were low in the original cohort (Table 1), all variables had a standardized difference less than 0. 1 after matching (Table 4). A univariate Cox proportional hazard model demonstrated that patients exposed to BBB ACEi had a 16% lower hazard of dementia compared to control cohort (HR 0.843, 95% CI 0.736-0.966; Table 5). A Kaplan Meir plot of dementia demonstrates that patients exposed to BBB ACEi had a higher probability of overall survival of dementia compared to the control BBB ACEi unexposed cohort (Figure 2, LogRank test = 0.0140). Since there are variables that may influence the development of dementia a multivariate Cox model was utilized to account for the baseline demographic and clinical characteristics (Table 6). After propensity score matching for BBB ACE inhibitor exposure, the baseline demographic and clinical characteristics between the two groups were well-balanced, with small, standardized differences and statistically significant differences observed in some variables, such as race, age, BMI, and Charlson comorbidity index. This matching approach strengthens the comparability between the groups and the result was consistent with the initial univariate and multivariate statistical models, exposure to BBB ACEi had a 15% lower hazard of incident dementia compared to BBB ACEi unexposed (HR 0.852, 95% CI 0.743-0.977).

**Figure 2:**
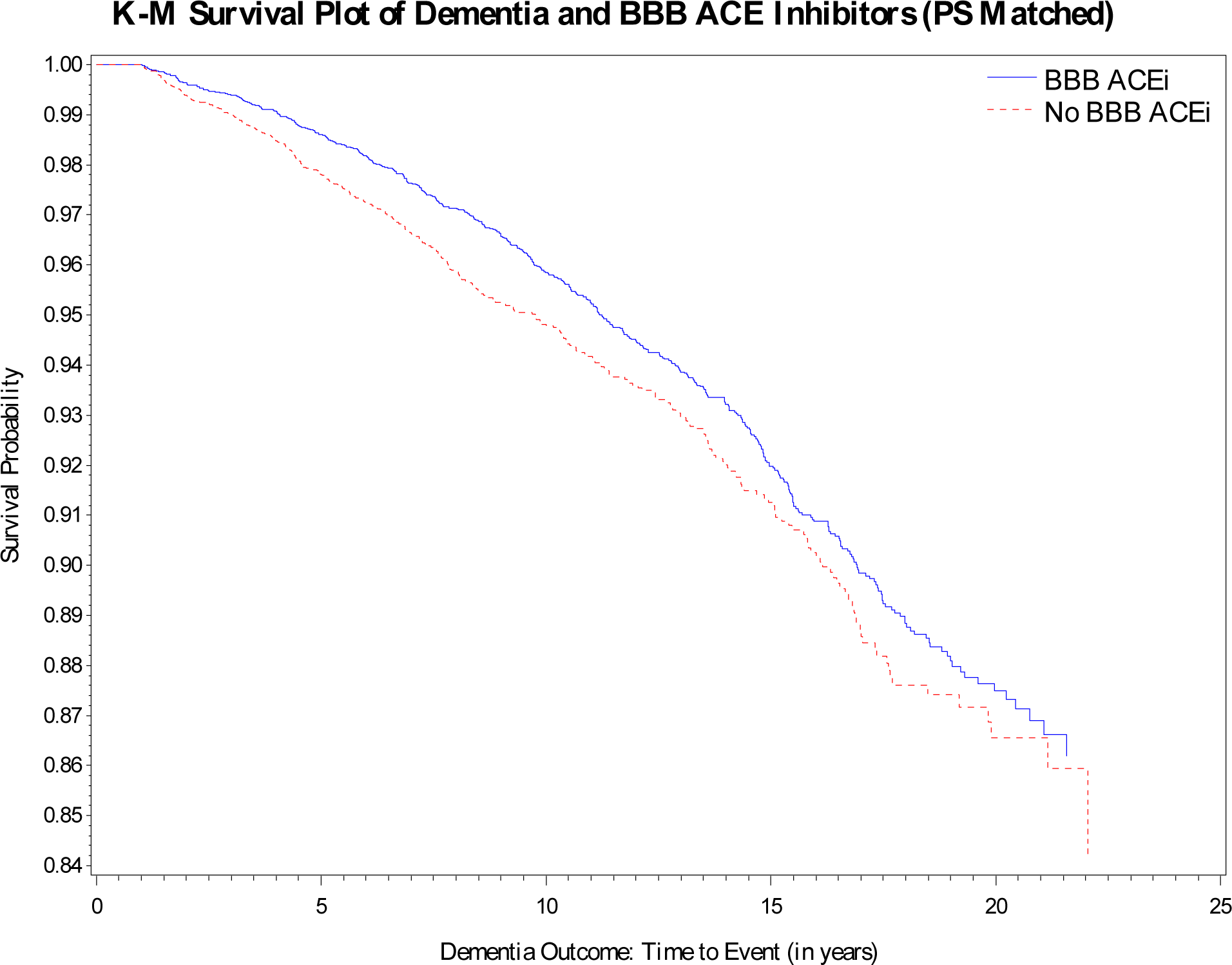
KM curve of propensity score model.

**Table 4:** Baseline Demographic and Clinical Characteristics for Propensity Score Matched Sample.

|  |  | BBB ACE Inhibitor<br>Exposed(N=7797) | BBB ACE<br>Inhibitor<br>Unexposed<br>(N=7797) | p-value | Standardized<br>Difference |
| --- | --- | --- | --- | --- | --- |
| Sex, No (%) | Female | 205(2.63) | 242(3.1) | 0.076 | 0.028 |
|  | Male | 7592(97.37) | 7555(96.9) |  | 0.028 |
| Race, No (%) | Black | 3925(50.34) | 3873(49.67) | 0.088 | 0.013 |
|  | Other/Unknown | 482(6.18) | 550(7.05) |  | 0.035 |
|  | White | 3390(43.48) | 3374(43.27) |  | 0.004 |
| Age, mean(SD) |  | 52.17(9.65) | 52.64(9.9) | 0.002 | 0.049 |
| Charlson comorbidity, mean(SD) |  | 4.77(3.11) | 5.05(3) | <0.001 | 0.091 |
| BMI, No (%) | <18.5 | 172(2.21) | 208(2.67) | 0.004 | 0.030 |
|  | 18.5-24.9 | 2708(34.73) | 2865(36.74) |  | 0.042 |
|  | 25.0-29.9 | 2945(37.77) | 2897(37.16) |  | 0.013 |
|  | 30+ | 1957(25.1) | 1806(23.16) |  | 0.045 |
|  | Missing | 15(0.19) | 21(0.27) |  | 0.017 |
| Pure hypercholesterolemia, No (%) |  | 204(2.62) | 198(2.54) | 0.762 | 0.005 |
| Hypertriglyceridemia, No (%) |  | 304(3.9) | 323(4.14) | 0.439 | 0.012 |
| Hyperlipidemia, No (%) |  | 1159(14.86) | 1296(16.62) | 0.003 | 0.048 |
| Ischemic heart disease, No (%) |  | 333(4.27) | 325(4.17) | 0.750 | 0.005 |
| Other heart disease, No (%) |  | 342(4.39) | 355(4.55) | 0.614 | 0.008 |
| T2DM, No (%) |  | 419(5.37) | 285(3.66) | <0.001 | 0.082 |
| Cerebral infarction, No (%) |  | 14(0.18) | 13(0.17) | 0.847 | 0.002 |
| Atrial fibrillation, No (%) |  | 31(0.4) | 35(0.45) | 0.622 | 0.008 |
| Hypothyroidism, No (%) |  | 85(1.09) | 100(1.28) | 0.267 | 0.018 |
| Hyperthyroidism, No (%) |  | 20(0.26) | 23(0.29) | 0.647 | 0.006 |
| Depression, No (%) |  | 1316(16.88) | 1312(16.83) | 0.932 | 0.001 |
| TBI, No (%) |  | 21(0.27) | 22(0.28) | 0.879 | 0.002 |
| Alcohol dependence, No (%) |  | 646(8.29) | 635(8.14) | 0.748 | 0.006 |
| Parkinson's disease, No (%) |  | ss | ss | 0.479 | 0.009 |
| Generalized anxiety disorder, No (%) |  | 101(1.3) | 105(1.35) | 0.779 | 0.004 |
| Chronic kidney disease, No (%) |  | 148(1.9) | 149(1.91) | 0.953 | 0.001 |
| Smoker, No (%) |  | 3788(48.58) | 3596(46.12) | 0.002 | 0.049 |
| Index Year, mean(SD) |  | 2004.67(5.64) | 2005.16(5.9) | <0.001 | 0.085 |

ss= sample size 10 patients or less.

**Table 5:** Univariate Results for Propensity Score Matched Sample.

|  |  | Cox Model |
| --- | --- | --- |
|  |  | HR (95% CI) |
| BBB ACE Inhibitor Exposure | Yes vs. No | 0.843 (0.736-0.966) |

**Table 6:** Multivariate Statistical Model for Incident Dementia for Propensity Score Matched Sample.

|  |  | Cox Model<br>HR (95% CI) |
| --- | --- | --- |
| BBB ACE Inhibitor Exposure | Yes vs. No | 0.852 (0.743-0.977) |
| Sex | Male vs. Female | 0.868 (0.565-1.334) |
| Race | Black vs. White | 1.415 (1.22-1.642) |
|  | Other/Unknown vs. White | 1.064 (0.76-1.489) |
| Age |  | 1.073 (1.064-1.082) |
| Charlson comorbidity |  | 1.008 (0.981-1.036) |
| BMI | <18.5 vs. 18.5-24.9 | 1.284 (0.873-1.89) |
|  | 25.0-29.9 vs. 18.5-24.9 | 0.853 (0.731-0.996) |
|  | 30+ vs. 18.5-24.9 | 0.823 (0.678-1) |
|  | Missing vs. 18.5-24.9 | 0 (0-1.93E+110) |
| Pure hypercholesterolemia |  | 0.688 (0.448-1.054) |
| Hypertriglyceridemia |  | 0.808 (0.551-1.183) |
| Hyperlipidemia |  | 0.994 (0.821-1.203) |
| Ischemic heart disease |  | 1.287 (0.959-1.729) |
| Other heart disease |  | 1.098 (0.805-1.496) |
| T2DM |  | 1.423 (1.09-1.856) |
| Cerebral infarction |  | 2.755 (1.021-7.435) |
| Atrial fibrillation |  | 1.218 (0.547-2.71) |
| Hypothyroidism |  | 1.286 (0.789-2.097) |
| Hyperthyroidism |  | 0.617 (0.152-2.5) |
| Depression |  | 1.39 (1.172-1.649) |
| TBI |  | 2.044 (0.762-5.483) |
| Alcohol dependence |  | 1.143 (0.908-1.437) |
| Parkinson's disease |  | 11.724 (4.252-32.327) |
| Generalized anxiety disorder |  | 0.854 (0.475-1.537) |
| Chronic kidney disease |  | 1.12 (0.708-1.773) |
| Smoker |  | 1.005 (0.87-1.161) |
| Index Year |  | 0.987 (0.97-1.004) |
| E-values: Point Estimate |  | 1.63 |
| Confidence Interval |  | 1.18 |

There are limitations to our drug disease observational study intrinsic to all health insurance claims database analyses, particularly proper documentation and coding. The study utilized data from the Veterans Affairs Informatics and Computing Infrastructure (VINCI) and did not utilize registries, other data sets, or potential data from the Veterans Choice Program. An additional study limitation is the population was United States Veterans; therefore, our findings may not be generalizable to patients of different age groups or races. Specifically, the cohort of patients may have different proportions of comorbidities, and although we controlled for many comorbidities within our statistical models, the results may not be reflective of non–Veterans Affairs populations. As with all retrospective studies, including ours, treatment was not randomized and differences among the treatment groups could influence the outcome. Therefore, we conducted a sub-analysis utilizing propensity score matching to assemble cohorts of patients with similar baseline characteristics with the attempt to reduce possible bias in estimating the treatment effect. To further explore our findings, we conducted an additional sub-analysis comparing PLWH and BBB ACEi to PLWH and non-BBB ACEI. BBB ACE inhibitors consisted of captopril, fosinopril, lisinopril, perindopril, ramipril, and trandolapril and non-BBB ACE inhibitors included: benazepril, enalapril, moexipril, and quinapril. In this sub-analysis, a total of 9,111 ACE inhibitor exposed patients met the inclusion and exclusion criteria. The sub cohorts consisted of 8,768 patients BBB ACE inhibitor exposed compared to 343 patients exposed to a non-BBB ACE inhibitor (Supplemental Table 1). The univariate Cox proportional hazards model demonstrated that patients exposed to BBB ACEi had a 21% lower hazard of dementia compared to non-BBB ACEi cohort (HR 0.786, 95% CI 0.508-1.216; Supplemental Table 2). After propensity score matching, the exposure groups are extremely well-balanced (Supplemental Table 3) and patients exposed to BBB ACEi had a 52.6% lower hazard of dementia compared to non-BBB ACE I cohort (0.474, 95% CI 0.228-0.984; Supplemental Table 4).

## 4.0 Discussion and Conclusion

We report application of an AI-based literature mining system to uncover drugs with potential to be repurposed for HAND. In support of this, we report a retrospective cohort drug-disease association study to explore the impact of an already approved therapy for a different indication, hypertension, on a different disease outcome, dementia, in PLWH. Our analysis compared individuals exposed to BBB-penetrable ACE inhibitors with those unexposed to BBB penetrable ACE inhibitors. The findings revealed that individuals exposed to BBB ACE inhibitors had a lower hazard of dementia compared to those without exposure to BBB ACE inhibitors. PLWH patients not prescribed BBB ACE inhibitors served as a control for ACEi exposure and their general anti-hypertensive and other potential systemic effects. These results suggest there is a potential protective effect of BBB ACE inhibitor exposure against dementia in PLWH that warrants further investigation. To mitigate the inherent limitations of retrospective analyses of EHRs, propensity score was utilized to assemble matching cohorts of patients with similar baseline characteristics to reduce possible bias in estimating treatment effects. The baseline demographic and clinical characteristics between the two groups were well-balanced, with small, standardized differences and statistically significant differences observed in some variables, such as race, age, BMI, and Charlson comorbidity index. This matching approach strengthens the comparability between the groups and the result was consistent with the initial univariate and multivariate statistical models. Nonetheless, other limitations remain, for example the inability to categorize the severity of the dementia.

The impetus for this study stemmed from findings from our AI-based literature mining systems and validate the generalized approach of coupling AI-based biomedical literature mining to retrospective analyses of EHRs to determine potential associations between diseases and novel treatment options. There is emerging evidence from studies of other neurodegenerative conditions that patients receiving antihypertensives have a decreased risk of dementia and ACE inhibitors were reported to confer the greatest benefit, but published results thus far have not agreed.^7–10,12,13,17,18^ Our study specifically compared BBB-penetrable ACE inhibitors with non-penetrable counterparts to explore the connection more thoroughly. The advantages of our study are that it was performed in a defined patient population with an easily identifiable diagnosis in EHRs (PLWH) and for whom the elevated risk of HAND and dementia is known. In the biomedical literature, there are growing connections between the RAAS system and neurocognitive disorders, chiefly in models of Alzheimers^33–36^, but to our knowledge this is the first report of a connection to dementia in PWLH. Future studies are planned to explore the mechanistic underpinnings of these observations in vitro and in vivo, as well as further analysis of EHRs to better understand the depth of the effect regarding timing and duration of BBB ACE exposure.

Overall, our results strongly suggest that BBB-penetrating ACE inhibitors are beneficial in preventing cognitive decline in PLWH. Therefore, an adjustment of prescription recommendation of ACEi towards BBB-penetrating drugs for the treatment of hypertension and other conditions to PLWH may provide high impact on the quality of life by delaying age and HIV-related dementia.

## Data Availability

These analyses were performed using data that are available only within the US Department of Veterans Affairs secure research environment, the VA Informatics and Computing Infrastructure (VINCI). All relevant data outputs are within the paper and its supplemental information. All relevant output data in the present work are contained in the manuscript.

## Acknowledgements

The content of this article is solely the responsibility of the authors and does not necessarily represent the official views of the US Department of Veterans Affairs, nor does mention of trade names, commercial products or organizations imply endorsement by the US government. This paper represents, in part, original research conducted using data from the Department of Veterans Affairs and is the result of work supported with resources and the use of facilities at the Dorn Research Institute, Columbia VA Health Care System, Columbia, South Carolina. The National Institutes of Health is acknowledged for funding NIH/NIDA R01 DA054992 to MS, MDW, SSS, JM, TC, IS.

## Supplemental Material

**Supplemental Table 1:** Baseline Demographic and Clinical Characteristics.

|  |  | BBB ACE Inhibitor<br>Exposure<br>(N=8768) | Non-BBB ACE<br>Inhibitor<br>Exposure<br>(N=343) | p-value | Standardized<br>Difference |
| --- | --- | --- | --- | --- | --- |
| Sex, No (%) | Female | 217(2.47) | ss | 0.394 | 0.050 |
|  | Male | 8551(97.53) | 337(98.25) |  | 0.050 |
| Race, No (%) | Black | 4446(50.71) | 157(45.77) | 0.013 | 0.099 |
|  | Other/Unknown | 488(5.57) | 31(9.04) |  | 0.134 |
|  | White | 3834(43.73) | 155(45.19) |  | 0.029 |
| Age, mean(SD) |  | 52.04(9.47) | 53.39(8.93) | 0.009 | 0.147 |
| Charlson comorbidity, mean(SD) |  | 4.71(3.13) | 4.03(3.41) | <0.001 | 0.209 |
| BMI, No (%) | <18.5 | 177(2.02) | ss | 0.044 | 0.043 |
|  | 18.5-24.9 | 2984(34.03) | 121(35.28) |  | 0.026 |
|  | 25.0-29.9 | 3381(38.56) | 130(37.9) |  | 0.014 |
|  | 30+ | 2212(25.23) | 84(24.49) |  | 0.017 |
|  | Missing | 14(0.16) | ss |  | 0.099 |
| Pure hypercholesterolemia, No (%) |  | 235(2.68) | 12(3.5) | 0.360 | 0.047 |
| Hypertriglyceridemia, No (%) |  | 354(4.04) | ss | 0.016 | 0.158 |
| Hyperlipidemia, No (%) |  | 1242(14.17) | 50(14.58) | 0.830 | 0.012 |
| Ischemic heart disease, No (%) |  | 358(4.08) | 16(4.66) | 0.594 | 0.028 |
| Other heart disease, No (%) |  | 359(4.09) | ss | 0.104 | 0.100 |
| T2DM, No (%) |  | 595(6.79) | 22(6.41) | 0.788 | 0.015 |
| Cerebral infarction, No (%) |  | 13(0.15) | ss | 0.506 | 0.030 |
| Atrial fibrillation, No (%) |  | 36(0.41) | ss | 0.734 | 0.020 |
| Hypothyroidism, No (%) |  | 82(0.94) | ss | 0.218 | 0.083 |
| Hyperthyroidism, No (%) |  | 20(0.23) | ss | 0.810 | 0.012 |
| Depression, No (%) |  | 1415(16.14) | 52(15.16) | 0.629 | 0.027 |
| TBI, No (%) |  | 20(0.23) | ss | 0.810 | 0.012 |
| Alcohol dependence, No (%) |  | 704(8.03) | 21(6.12) | 0.201 | 0.075 |
| Parkinson's disease, No (%) |  | ss | 0(0) | 0.732 |  |
| Generalized anxiety disorder, No (%) |  | 104(1.19) | ss | 0.349 | 0.047 |
| Chronic kidney disease, No (%) |  | 156(1.78) | ss | 0.720 | 0.019 |
| Smoker, No (%) |  | 4294(48.97) | 161(46.94) | 0.460 | 0.041 |
| Index Year, mean(SD) |  | 2004.39(5.52) | 2006.16(6.11) | <0.001 | 0.304 |

ss= sample size is 10 patients or less.

**Supplemental Table 2:** Univariate Results.

|  |  | Cox Model |
| --- | --- | --- |
|  |  | HR (95% CI) |
| BBB ACE Inhibitor Exposure | BBB ACEi vs. Non-BBB ACEi | 0.786 (0.508-1.216) |

**Supplemental Table 3:** Baseline Demographic and Clinical Characteristics for Propensity Score Matched Sample.

|  |  | BBB ACE Inhibitor<br>Exposure (N=340) | Non-BBB ACE<br>Inhibitor<br>Exposure<br>(N=340) | p-value | Standardized<br>Difference |
| --- | --- | --- | --- | --- | --- |
| Sex, No (%) | Female | ss | ss | 0.314 | 0.077 |
|  | Male | 337(99.12) | 334(98.24) |  | 0.077 |
| Race, No (%) | Black | 156(45.88) | 156(45.88) | 1.000 | 0.000 |
|  | Other/Unknown | 30(8.82) | 30(8.82) |  | 0.000 |
|  | White | 154(45.29) | 154(45.29) |  | 0.000 |
| Age, mean(SD) |  | 53.36(8.74) | 53.28(8.88) | 0.896 | 0.010 |
| Charlson comorbidity, mean(SD) |  | 3.54(3.36) | 4(3.41) | 0.078 | 0.135 |
| BMI, No (%) | <18.5 | ss | ss | 1.000 | 0.000 |
|  | 18.5-24.9 | 121(35.59) | 121(35.59) |  | 0.000 |
|  | 25.0-29.9 | 130(38.24) | 130(38.24) |  | 0.000 |
|  | 30+ | 84(24.71) | 84(24.71) |  | 0.000 |
|  | Missing | ss | ss |  | 0.000 |
| Pure hypercholesterolemia, No (%) |  | 13(3.82) | 11(3.24) | 0.678 | 0.031 |
| Hypertriglyceridemia, No (%) |  | ss | ss | 1.000 | 0.000 |
| Hyperlipidemia, No (%) |  | 47(13.82) | 49(14.41) | 0.826 | 0.017 |
| Ischemic heart disease, No (%) |  | ss | 15(4.41) | 0.308 | 0.078 |
| Other heart disease, No (%) |  | ss | ss | 0.560 | 0.045 |
| T2DM, No (%) |  | 26(7.65) | 22(6.47) | 0.549 | 0.046 |
| Cerebral infarction, No (%) |  | ss | ss | 0.563 | 0.045 |
| Atrial fibrillation, No (%) |  | 0(0) | ss | 0.317 |  |
| Hypothyroidism, No (%) |  | ss | ss | 1.000 | 0.000 |
| Hyperthyroidism, No (%) |  | ss | ss | 0.563 | 0.045 |
| Depression, No (%) |  | 36(10.59) | 52(15.29) | 0.068 | 0.140 |
| TBI, No (%) |  | 0(0) | ss | 0.317 |  |
| Alcohol dependence, No (%) |  | 22(6.47) | 21(6.18) | 0.875 | 0.012 |
| Parkinson's disease, No (%) |  | 0(0) | 0(0) |  |  |
| Generalized anxiety disorder, No (%) |  | ss | ss | 0.779 | 0.022 |
| Chronic kidney disease, No (%) |  | ss | ss | 0.361 | 0.070 |
| Smoker, No (%) |  | 152(44.71) | 160(47.06) | 0.538 | 0.047 |
| Index Year, mean(SD) |  | 2006.27(6.29) | 2006.16(6.06) | 0.804 | 0.019 |

ss= sample size is 10 patients or less.

**Supplemental Table 4:** Univariate Results for Propensity Score Matched Sample.

|  |  | Cox Model |
| --- | --- | --- |
|  |  | HR (95% CI) |
| BBB ACE Inhibitor Exposure | BBB ACEi vs. Non-BBB ACEi | 0.474 (0.228-0.984) |

## References

1. Yoshimura, K. (2017). Current status of HIV/AIDS in the ART era. J. Infect. Chemother. 23.

2. Osborne, O., Peyravian, N., Nair, M., Daunert, S., and Toborek, M. (2020). The Paradox of HIV Blood–Brain Barrier Penetrance and Antiretroviral Drug Delivery Deficiencies. Trends Neurosci. 43.

3. CDC (2020). HIV in the United States by Age. https://www.cdc.gov/hiv/group/age/index.html.

4. Smit, M., Brinkman, K., Geerlings, S., Smit, C., Thyagarajan, K., van Sighem, A. V., de Wolf, F., and Hallett, T.B. (2015). Future challenges for clinical care of an ageing population infected with HIV: A modelling study. Lancet Infect. Dis. 15.

5. Sybrandt, J., Shtutman, M., and Safro, I. (2017). MOLIERE: Automatic biomedical hypothesis generation system. In Proceedings of the ACM SIGKDD International Conference on Knowledge Discovery and Data Mining.

6. Sybrandt, J., Shtutman, M., and Safro, I. (2019). Large-Scale Validation of Hypothesis Generation Systems via Candidate Ranking. In Proceedings - 2018 IEEE International Conference on Big Data, Big Data 2018.

7. Aksenova, M., Sybrandt, J., Cui, B., Sikirzhytski, V., Ji, H., Odhiambo, D., Lucius, M.D., Turner, J.R., Broude, E., Peña, E., et al. (2020). Inhibition of the Dead Box RNA Helicase 3 Prevents HIV-1 Tat and Cocaine-Induced Neurotoxicity by Targeting Microglia Activation. J. Neuroimmune Pharmacol. 15.

8. Sybrandt, J., Tyagin, I., Shtutman, M., and Safro, I. (2020). AGATHA: Automatic Graph Mining and Transformer based Hypothesis Generation Approach. In International Conference on Information and Knowledge Management, Proceedings.

9. Cui BC, Aksenova M, Sikirzhytskaya A, Odhiambo D, Korunova E, Sikirzhytski V, Ji H, Altomare D, Broude E, Frizzell N, Booze R, Wyatt MD, S.M. (2023). Suppression of HIV and cocaine-induced neurotoxicity and inflammation by cell penetrable itaconate esters. BioRxiv 2023.09.25.

10. Nehme, A., and Zibara, K. (2017). Efficiency and specificity of RAAS inhibitors in cardiovascular diseases: How to achieve better end-organ protection? Hypertens. Res. 40.

11. Simmons, M., Singhal, A., and Lu, Z. (2016). Text mining for precision medicine: Bringing structure to ehrs and biomedical literature to understand genes and health. In Advances in Experimental Medicine and Biology.

12. Sink, K.M., Leng, X., Williamson, J., Kritchevsky, S.B., Yaffe, K., Kuller, L., Yasar, S., Atkinson, H., Robbins, M., Psaty, B., et al. (2009). Angiotensin-converting enzyme inhibitors and cognitive decline in older adults with hypertension: Results from the cardiovascular health study. Arch. Intern. Med. 169.

13. Ho, J.K., Moriarty, F., Manly, J.J., Larson, E.B., Evans, D.A., Rajan, K.B., Hudak, E.M., Hassan, L., Liu, E., Sato, N., et al. (2021). Blood-Brain Barrier Crossing Renin-Angiotensin Drugs and Cognition in the Elderly: A Meta-Analysis. Hypertension 78.

14. Van Der Weele, T.J., and Ding, P. (2017). Sensitivity analysis in observational research: Introducing the E-Value. Ann. Intern. Med. 167, 268–74.

15. Mathur, M.B., Ding, P., Riddell, C.A., and VanderWeele, T.J. (2018). Website and R Package for Computing E-values. Epidemiology 29, e45–e47.

16. Cui, B.C., Sikirzhytski, V., Aksenova, M., Lucius, M.D., Levon, G.H., Mack, Z.T., Pollack, C., Odhiambo, D., Broude, E., Lizarraga, S.B., et al. (2020). Pharmacological inhibition of DEAD-Box RNA Helicase 3 attenuates stress granule assembly. Biochem. Pharmacol. 182.

17. Laghlam, D., Jozwiak, M., and Nguyen, L.S. (2021). Renin–angiotensin–aldosterone system and immunomodulation: A state-of-the-art review. Cells 10.

18. Arroja, M.M.C., Reid, E., and McCabe, C. (2016). Therapeutic potential of the renin angiotensin system in ischaemic stroke. Exp. Transl. Stroke Med. 8.

19. Fazal, K., Perera, G., Khondoker, M., Howard, R., and Stewart, R. (2017). Associations of centrally acting ACE inhibitors with cognitive decline and survival in Alzheimer’s disease. BJPsych Open 3.

20. Duron, E., and Hanon, O. (2008). Hypertension, cognitive decline and dementia. Arch. Cardiovasc. Dis. 101, 181–189.

21. de la Torre, J.C. (2002). Alzheimer Disease as a Vascular Disorder. Stroke 33.

22. Zlokovic, B. V. (2011). Neurovascular pathways to neurodegeneration in Alzheimer’s disease and other disorders. Nat. Rev. Neurosci. 12.

23. Saxby, B.K., Harrington, F., McKeith, I.G., Wesnes, K., and Ford, G.A. (2003). Effects of Hypertension on Attention, Memory, and Executive Function in Older Adults. Heal. Psychol. 22.

24. Waldstein, S.R. (2003). The relation of hypertension to cognitive function. Curr. Dir. Psychol. Sci. 12.

25. Levi Marpillat, N., MacQuin-Mavier, I., Tropeano, A.I., Bachoud-Levi, A.C., and Maison, P. (2013). Antihypertensive classes, cognitive decline and incidence of dementia: A network meta-analysis. J. Hypertens. 31.

26. Davies, N.M., Kehoe, P.G., Ben-Shlomo, Y., and Martin, R.M. (2011). Associations of anti-hypertensive treatments with Alzheimer’s disease, vascular dementia, and other dementias. J. Alzheimer’s Dis. 26.

27. Barthold, D., Joyce, G., Wharton, W., Kehoe, P., and Zissimopoulos, J. (2018). The association of multiple anti-hypertensive medication classes with Alzheimer’s disease incidence across sex, race, and ethnicity. PLoS One 13.

28. Ye, R., Hu, Y., Yao, A., Yang, Y., Shi, Y., Jiang, Y., and Zhang, J. (2015). Impact of renin - angiotensin system-targeting antihypertensive drugs on treatment of Alzheimer’s disease: A meta-analysis. Int. J. Clin. Pract. 69.

29. Hajjar, I., Okafor, M., McDaniel, D., Obideen, M., Dee, E., Shokouhi, M., Quyyumi, A.A., Levey, A., and Goldstein, F. (2020). Effects of Candesartan vs Lisinopril on Neurocognitive Function in Older Adults with Executive Mild Cognitive Impairment: A Randomized Clinical Trial. JAMA Netw. Open 3.

30. McGuinness, B., Todd, S., Passmore, P., and Bullock, R. (2009). Blood pressure lowering in patients without prior cerebrovascular disease for prevention of cognitive impairment and dementia. Cochrane Database Syst. Rev.

31. Fournier, A., Oprisiu-Fournier, R., Serot, J.M., Godefroy, O., Achard, J.M., Faure, S., Mazouz, H., Temmar, M., Albu, A., Bordet, R., et al. (2009). Prevention of dementia by antihypertensive drugs: How AT1-receptor-blockers and dihydropyridines better prevent dementia in hypertensive patients than thiazides and ACE-inhibitors. Expert Rev. Neurother. 9.

32. Anderson, C., Teo, K., Gao, P., Arima, H., Dans, A., Unger, T., Commerford, P., Dyal, L., Schumacher, H., Pogue, J., et al. (2011). Renin-angiotensin system blockade and cognitive function in patients at high risk of cardiovascular disease: Analysis of data from the ONTARGET and TRANSCEND studies. Lancet Neurol. 10.

33. Ababei, D.C., Bild, V., Macadan, I., Vasincu, A., Rusu, R.N., Blaj, M., Stanciu, G.D., Lefter, R.M., and Bild, W. (2023). Therapeutic Implications of Renin–Angiotensin System Modulators in Alzheimer’s Dementia. Pharmaceutics 15.

34. Yang, M.H., Ho, T.C., Chang, C.C., Su, Y.S., Yuan, C.H., Chuang, K.P., and Tyan, Y.C. (2023). Utilizing Proteomic Approaches to Uncover the Neuroprotective Effects of ACE Inhibitors: Implications for Alzheimer’s Disease Treatment. Molecules 28.

35. Thomas, J., Smith, H., Smith, C.A., Coward, L., Gorman, G., De Luca, M., and Jumbo-Lucioni, P. (2021). The angiotensin-converting enzyme inhibitor lisinopril mitigates memory and motor deficits in a drosophila model of alzheimer’s disease. Pathophysiology 28.

36. Cuddy, L.K., Prokopenko, D., Cunningham, E.P., Brimberry, R., Song, P., Kirchner, R., Chapman, B.A., Hofmann, O., Hide, W., Procissi, D., et al. (2020). Aβ-accelerated neurodegeneration caused by Alzheimer’s-associated ACE variant R1279Q is rescued by angiotensin system inhibition in mice. Sci. Transl. Med. 12.

